# Ecological Analysis of the Temporal Trends in the Association of Social Vulnerability and Race/Ethnicity with County-Level COVID-19 Incidence and Outcomes in the United States

**DOI:** 10.1101/2021.06.04.21258355

**Authors:** Shabatun J. Islam, Aditi Nayak, Yingtian Hu, Anurag Mehta, Katherine Dieppa, Zakaria Almuwaqqat, Yi-An Ko, Shivani A. Patel, Abhinav Goyal, Samaah Sullivan, Tené T. Lewis, Viola Vaccarino, Alanna A. Morris, Arshed A. Quyyumi

## Abstract

**Background:** The COVID-19 pandemic adversely affected the socially vulnerable and minority communities in the U.S. initially, but the temporal trends during the year-long pandemic remain unknown.

**Objective:** We examined the temporal association between the county-level Social Vulnerability Index (SVI), a percentile-based measure of social vulnerability to disasters, its subcomponents and race/ethnic composition with COVID-19 incidence and mortality in the U.S. in the year starting in March 2020.

**Methods:** Counties (n=3091) with ≥ 50 COVID-19 cases by March 6^th^, 2021 were included in the study. Associations between SVI (and its subcomponents) and county level racial composition with the incidence and death per capita were assessed by fitting a negative-binomial mixed-effects model. This model was also used to examine potential time varying associations between weekly number of cases/deaths and SVI or racial composition. Data was adjusted for percentage of population aged ≥65 years, state level testing rate, comorbidities using the average Hierarchical Condition Category (HCC) score, and environmental factors including average fine particulate matter (PM_2.5_), temperature and precipitation.

**Results:** Higher SVI, indicative of greater social vulnerability, was independently associated with higher COVID-19 incidence (adjusted incidence rate ratio [IRR] per-10 percentile increase:1.02, (95% CI 1.02, 1.03, p<0.001), and death per capita (1.04, (95% CI 1.04, 1.05, p<0.001). SVI became an independent predictor of incidence starting from March 2020, but this association became weak or insignificant by the winter, a period that coincided with a sharp increase in infection rates and mortality, and when counties with higher proportion of White residents were disproportionately represented (“third wave”). By Spring of 2021, SVI was again a predictor of COVID-19 outcomes. Counties with greater proportion of Black residents also observed similar temporal trends COVID-19-related adverse outcomes. Counties with greater proportion of Hispanic residents had worse outcomes throughout the duration of the analysis.

**Conclusion:** Except for the winter “third wave” when majority White communities had the highest incidence of cases, counties with greater social vulnerability and proportionately higher minority populations, experienced worse COVID-19 outcomes.

**Article Summary/Strengths & Limitations:**

- Examined full 12 months of county-level data in the US delineating the temporal trends in the association between social vulnerability index and COVID-19 outcomes
- Investigated COVID-19 outcomes in predominantly Black and Hispanic communities in comparison to White communities in the US
- Analysis is ecological, descriptive, and on the county-level rather than on an individual level
- Analysis adjusted for confounders including county level age ≥ 65, comorbidities, and environmental factors
- Analysis limited to the US

## Introduction

Community-level social disadvantage and vulnerability to disasters, as well as race/ethnic composition can influence the incidence of COVID-19 and its adverse outcomes in several ways. For example, lower socioeconomic status (SES) is associated with uncertain healthcare access, poor health status and higher risk factor burden that together contribute to a greater risk of adverse outcomes.^1^ Labor inequalities and household overcrowding may decrease the ability to adhere to social-distancing guidelines.^2^ Black and Hispanic individuals are more likely to work in front-line jobs with lack of workplace protections that may additionally increase exposure risk.^3^ Additionally, race/ethnic minorities and immigrants are less likely to have access to appropriate and timely healthcare.^3–5^ Evidence suggests that these inequalities also contributed to disease spread and adverse outcomes during the H1N1 influenza pandemic.^6,7^

The Social Vulnerability Index (SVI), created and maintained by the Geospatial Research, Analysis, and Services Program (GRASP) at the Centers for Disease Control and Prevention (CDC) and Agency for Toxic Substances and Disease Registry, is a percentile-based index of county-level vulnerability to disasters and was designed for resource allocation to vulnerable communities during times of duress such as the COVID-19 pandemic.^8,9^ The SVI includes measures of county-level socioeconomic status, housing composition and disability, minority status and language, and housing type and transport, and thus allows for a dynamic understanding of challenges encountered by communities. Emerging data during the COVID-19 pandemic has demonstrated that socially vulnerable neighborhoods have had worse outcomes during the early stages of the pandemic,^10–14^ even given the fact that the SVI had been designed to mitigate such adverse outcomes for vulnerable communities. Data has shown that the association between SVI and COVID-19 outcomes temporally varied, with the trend reversing by October 2020,^15^ but whether this continued to the latter durations of the pandemic in unknown.

Similarly, it is now well known that Black^16,17^ and Hispanic^18^ individuals, who represent the largest minority groups within the U.S., are especially susceptible to worse COVID-19 outcomes, but the temporal trend of these associations throughout the course of the pandemic remains unknown. Herein, we first report on the temporal trends in the association of county-level SVI and its subcomponents with COVID-19 incidence and death per capita in the U.S. for the entire year from March 2020 to March 2021. Secondly, since the SVI subcomponent of minority status and language does not delineate specific racial ethnic composition, we also examine the temporal trends of the association of county-level proportion of Black and Hispanic residents and COVID-19 outcomes.

## Methods

### Study population & time frame

All U.S. counties (n=3091) with at least 50 confirmed COVID-19 cases and greater than 4-week of follow-up data were included in the analysis. Data was analyzed for a period of 50 weeks starting from March 22, 2020 to March 6^th^, 2021.

### Patient and Public Involvement Statement

The patient and the public were not involved in the design of this study.

### Outcomes

Primary outcomes of interest were county-level weekly COVID-19 incidence and death per capita of a county. Data were obtained from the Johns Hopkins Center for Systems Science and Engineering database.^1^

### Exposures

Exposures studied were (a) 2018 county-level SVI and its subcomponents obtained from the CDC GRASP database,^2,3^ and (b) racial composition data (reported as proportion of Black and Hispanic residents in a county) from the U.S. Census Bureau ACS database.^4^ The SVI was developed by the CDC as a measure of community resilience to stresses on human health such as disease outbreaks and natural or human-caused disasters, to help public health officials and emergency response planners identify communities that are likely to need support before, during, and after a disaster. ^2,3^ The index combines statistical data from the U.S. Census on 15 variables, grouped together into four related themes: socioeconomic status (SES), housing composition and disability, minority status and language, and housing type and transport (**Table 1**). Each of these variables are ranked from lowest to highest vulnerability across census tracts in the U.S. and a county-level percentile rank is calculated for each variable, theme, and the overall SVI, with higher percentiles indicating higher social vulnerability. In terms of the racial composition data, we focused on the proportion of Black and Hispanic residents in a county since they represent the largest minority groups across a broader geographic region in the U.S.. The data was collected by the U.S. Census Bureau as self-reported race/ethnicity between 2015-2019. ^4^

**Table 1.** Components of the social vulnerability index

|  |  |
| --- | --- |
| **Socioeconomic Status** | Below Poverty<br>Unemployed<br>Income<br>No High School Diploma |
| **Household composition and Disability** | Age 65 years or older<br>Age 17 years or younger<br>Older than Age 5 years with a Disability |
| **Minority Status and Language** | Minority<br>Speak English "Less than Well" |
| **Housing Type and Transportation** | Multi-Unit Structures<br>Mobile Homes<br>Crowding<br>No Vehicle<br>Group Quarters |

### Confounders

Covariates included in all models were proportion of county population aged ≥65 years^4^, state-level COVID-19 testing rate obtained from the COVID Tracking Project database,^5^ 2018 Hierarchical Condition Category (HCC) risk score acquired from the Centers for Medicare and Medicaid Services (CMS) database as a proxy for county-level medical comorbidity, and environmental factors. State-level COVID-19 testing rate is calculated as all tests completed (whether symptomatic and asymptomatic, voluntary or contact tracing) divided by the state-level population. The HCC risk score, based on medical risk profiles and demographics of county Medicare beneficiaries, was developed by CMS to risk-adjust Medicare spending for beneficiary health status.^19,20^ While the score was designed to reflect healthcare access and hospital admissions in a geographic area, it does compare favorably to other comorbidity indices in prediction of outcomes,^19^ and aggregate county-level scores are publicly available.^20^ As such, we are using the HCC risk score as a proxy for county-level comorbidities. For environmental factors, we included average daily temperature (degrees Fahrenheit),^21^ average daily precipitation,^21^ and average particulate matter of diameter ≥ 2.5 micrometers (PM_2.5_).^22^ All data sources used in this analysis are publicly available and are listed in **Table S1**.

### Statistical analysis

The overall associations between SVI (and its subcomponents) with the cumulative outcome variables including incidence and death per capita were assessed by fitting a negative-binomial mixed-effects model accounting for SVI as fixed effects with county specific random intercepts. The time-varying associations between SVI (and its subcomponents) of a county with the weekly outcome variables were assessed by fitting a negative-binomial mixed-effects model with weekly total confirmed case numbers or weekly total death numbers as the outcome and county-specific random intercepts to account for overdispersion, correlation in the outcome within counties, and heterogeneity across counties. The fixed effects included SVI, time (in weeks), and the interaction between time and SVI. Time was expressed using natural cubic splines with 3 degrees of freedom to allow for nonlinear relationships. Similarly, overall associations and time-varying associations between country-specific White, Black and Hispanic race/ethnic composition and weekly outcome variables were evaluated using the same model by replacing SVI with the respective race/ethnic composition variable. Total population in each county was used as the offset in all models. We further adjusted for covariates including percentage aged ≥65 years, state-level testing rate, HCC risk score, average daily temperature (degrees Fahrenheit),^21^ average daily precipitation,^21^ and average particulate matter of diameter ≥ 2.5 micrometers (PM_2.5_)^22^ as described above. All analyses were performed using R, version 3.6.1 (R Foundation for Statistical Computing). All *P* values were 2-sided, with a significance level of 0.05.

## RESULTS

Among the 3142 counties in the U.S., 3091 (98.38%) had >50 confirmed COVID-19 cases as of March 6th, 2021 and at least 4-week follow up data; accounting for a total of 28,547,384 cases from 362,058,535 administered tests, and 517,733 deaths. The median SVI for counties included in this analysis was 0.44 [Range: 0.17-0.85]. The median county-level COVID-19 incidence was 90.7 per 1000 people [Range: 2.61 – 368.2] and death per capita was 1.64 per 1000 people [Range 0.00-7.89]. The median proportions of White, Black and Hispanic residents per county were 89.4% [Range: 3.9-99.9%], 2.4% [Range: 0.0-87.4%] and 4.1% [Range: 0.0-99.1%], respectively. Overall SVI correlated most strongly with the subcomponent of socioeconomic status and least with minority status and language (Supplemental Figure 1).

Proportion of Black residents correlated modestly (r=0.4) and proportion of Hispanic residents correlated slightly (r=0.2) with overall SVI (Supplemental Figure 1).

### Overall and Temporal Associations between SVI and COVID-19 incidence

The incidence of COVID-19 infections was significantly higher in counties with greater SVI or higher social vulnerability, (adjusted incidence rate ratio [IRR] per-10 percentile increase: 1.02 [95% CI 1.02, 1.03], p<0.001) after adjusting for aforementioned confounders as of March 6th, 2021. Among the SVI sub-components, indices of SES (adjusted IRR per 10-percentile increase: 1.02, [95% CI 1.01, 1.03], p<0.001), minority status and language (adjusted IRR per 10-percentile increase: 1.02 [95% CI 1.01, 1.02], p<0.001), and household composition and disability (adjusted IRR per 10-percentile increase: 1.01 [95% CI 1.01, 1.02], p<0.001) were independently associated with COVID-19 incidence **(Table 2)**.

**Table 2.** Overall association of county-level social vulnerability index (Incidence Risk Ratio [IRR] per-10 percentile increase) with incidence and death per capita of COVID-19 as of March 6th, 2021

|  | Model 1<sup>a</sup> |  | Model 2<sup>b</sup> |  | Model 3<sup>c</sup> |  |
| --- | --- | --- | --- | --- | --- | --- |
|  | IRR (95% CI) | P-value | IRR (95% CI) | P-value | IRR (95% CI) | P-value |
| **Incidence** |  |  |  |  |  |  |
| **Overall Social Vulnerability Index\*** | 1.03 (1.03, 1.04) | <0.001 | 1.02 (1.02, 1.03) | <0.001 | 1.02 (1.02, 1.03) | <0.001 |
| Socioeconomic Status | 1.02 (1.02, 1.03) | <0.001 | 1.02 (1.01, 1.02) | <0.001 | 1.02 (1.01, 1.02) | <0.001 |
| Minority Status & Language | 1.02 (1.01, 1.02) | <0.001 | 1.02 (1.01, 1.02) | <0.001 | 1.02 (1.01, 1.02) | <0.001 |
| Housing Type & Transport | 1.01 (1.00, 1.01) | 0.003 | 0.99 (0.99, 1.00) | 0.07 | 0.99 (0.99, 1.00) | 0.29 |
| Household Composition & Disability* | 1.02 (1.01, 1.02) | <0.001 | 1.01 (1.01, 1.02) | <0.001 | 1.01 (1.01, 1.02) | <0.001 |
| **Death Per Capita** |  |  |  |  |  |  |
| **Overall Social Vulnerability Index\*** | 1.07 (1.06, 1.08) | <0.001 | 1.05 (1.04, 1.06) | <0.001 | 1.04 (1.04, 1.05) | <0.001 |
| Socioeconomic Status | 1.07 (1.07, 1.08) | <0.001 | 1.05 (1.04, 1.06) | <0.001 | 1.05 (1.04, 1.05) | <0.001 |
| Minority Status & Language | 1.03 (1.02, 1.03) | <0.001 | 1.01 (1.00, 1.02) | 0.003 | 1.01 (1.00, 1.02) | 0.004 |
| Housing Type & Transport | 1.06 (1.05, 1.06) | <0.001 | 1.05 (1.04, 1.05) | <0.001 | 1.05 (1.04, 1.05) | <0.001 |
| Household Composition & Disability* | 1.04 (1.03, 1.05) | <0.001 | 1.02 (1.01, 1.02) | <0.001 | 1.02 (1.01, 1.02) | <0.001 |

<sup>a</sup> Base Model - adjusted for proportion of population age $\geq 65$ years and state-level COVID-19 testing
<sup>b</sup> Base Model + CMS average Hierarchical Condition Category (HCC) score (proxy for comorbidities)
<sup>c</sup> Base Model + CMS average Hierarchical Condition Category (HCC) score (proxy for comorbidities) + Environmental Factors including average daily temperature (degrees Fahrenheit), average daily precipitation, and average particulate matter of diameter $\geq 2.5$ micrometers (PM<sub>2.5</sub>)
\* Proportion age $\geq 65$ years not included as a covariate for models for overall social vulnerability index and household composition/disability because these indices contain the age variable.

#### Temporal Trends

**Figure 1** demonstrates the temporal trends in the incidence of infections in relation to the overall SVI and its components. As shown, overall county-level SVI was positively associated with COVID-19 incidence starting from the beginning of our analysis on March 22nd, 2020 (Week 1), with the association becoming stronger over time. However, the association weakened after mid-July, 2020 (Week 17) and there was no significant association between overall SVI and COVID-19 incidence between late October, 2020 (Week 32) and early December, 2020 (Week 37). This coincided with a large increase in cases within the U.S. (“third wave”). However, once the overall case load started to decrease from the peak by January, 2021 (Week 40) to early March 2021, overall SVI again demonstrated strong associations with COVID-19 incidence.

#### SVI Subcomponents (Figure 1)

**Figure 1:**
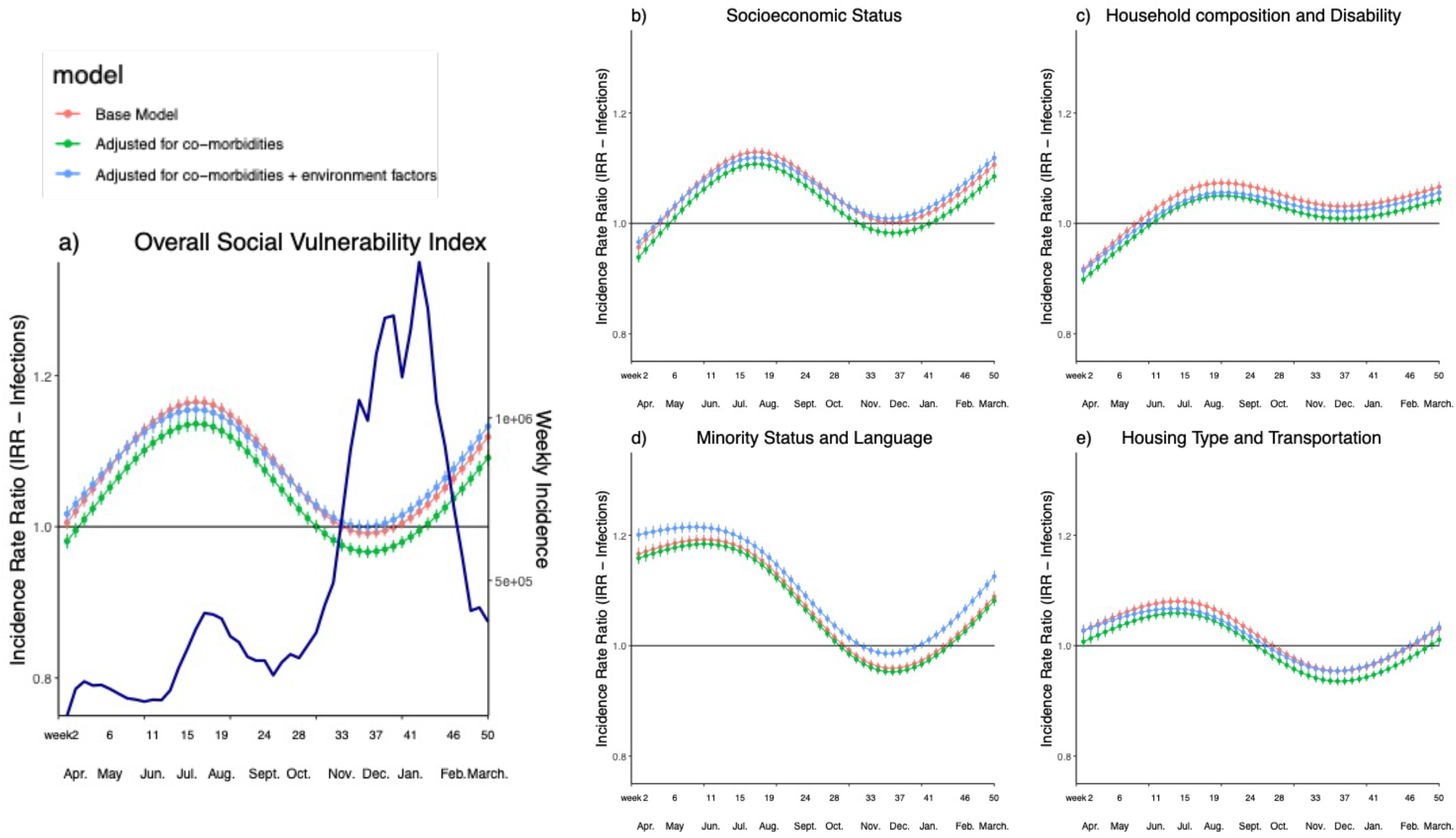
Temporal association between COVID-19 incidence and the a) county-level Social Vulnerability Index (SVI) and its subcomponents: b) Socioeconomic Status, c) Household Composition and Disability, d) Minority Status and Language, and e) Housing Type and Transportation between March 22^nd^, 2020 and March 6^th^, 2021. The base model (red lines) is adjusted for proportion of population age ≥65 years and state-level COVID-19 testing. The green lines are additionally adjusted for CMS average Hierarchical Condition Category (HCC) score (proxy for comorbidities). The blue lines are additionally adjusted for environmental factors including average daily temperature (degrees Fahrenheit), average daily precipitation, and average particulate matter of diameter ≥ 2.5 micrometers (PM_2.5_).Of note, proportion age ≥65 years not included as a covariate for models for overall social vulnerability index and household composition/disability because these indices contain the age variable.

The SVI subcomponent of minority status and language was an independent predictor of incidence from the beginning (March 22^nd^, 2020). While the association attenuated after adjustment for comorbidities using the HCC, it strengthened after additionally adjusting for environmental factors and remained positively associated with COVID-19 incidence until mid-October, 2020 (Week 30) when it started to be negatively associated with COVID-19 incidence with a rise in cases in the U.S (“third wave”). Similar to the overall SVI, it again became positively associated once the cases decreased in the U.S. around January-February 2021 (Week 41-46).

The SVI subcomponent of socioeconomic status was an independent predictor of incidence after accounting for co-morbidities and environmental factors starting in early May, 2020 (Week 6), with a strengthening association until mid-July, 2020 (Week 17), after which the association weakened and became insignificant by early November, 2020 (Week 33) but again became significant by mid December 2021 (Week 39).

The indices of county-level household composition and disability and housing type and transportation become independent predictors of incidence of COVID-19 in early June, 2020 (Week 11) and late March, 2020, (Week 1), respectively. The strength of the positive association varied for county-level household composition and disability but remained significant throughout the duration of our analysis. County-level housing type and transport remained a positive predictor of incidence until mid September 2020 (Week 26), became negative afterwards and became positive again in February 2021 (Week 48). Of note, in analysis additionally adjusting for percentage of residents under the federal poverty limit, for SVI subthemes of minority status and language, household composition and disability, and housing type and transport, similar trends are noted (Supplemental Figure 2).

### Overall and Temporal Associations between SVI and COVID-19 death per capita

The average death per capita from COVID-19 over the 50-week duration of the study was significantly higher in counties with greater SVI or higher social vulnerability (adjusted IRR per-10 percentile increase: 1.04, (95% CI [1.04, 1.05], p<0.001) after adjusting for aforementioned confounders (**Table 2**). All the SVI sub-components including indices of SES (adjusted IRR per 10-percentile increase: 1.05, 95% CI [1.04, 1.05], p<0.001), minority status and language (adjusted IRR per 10-percentile increase: 1.01, 95% CI [1.00, 1.02], p=0.004), housing type and transportation (adjusted IRR per 10-percentile increase: 1.05, 95% CI [1.04, 1.05], p<0.001), and household composition and disability (adjusted IRR per 10-percentile increase: 1.02, 95% CI [1.01, 1.02], p<0.001) were independently associated with COVID-19 death per capita (**Table 2**).

#### Temporal Trends

**Figure 2** demonstrates the temporal trends in death per capita in relation to the overall SVI and its components. As shown, overall county-level SVI firstly was an independent predictor of COVID-19 death per capita starting in early May, 2020 (Week 6) and the association became stronger over time. However, the association weakened after July, 2020 (Week 17) and became insignificant between November 2020 – January 2021 (Week 33 – 40) when cumulative deaths were at their highest. The association became significant starting in early January 2021 (Week 41) once the deaths started decreasing.

**Figure 2:**
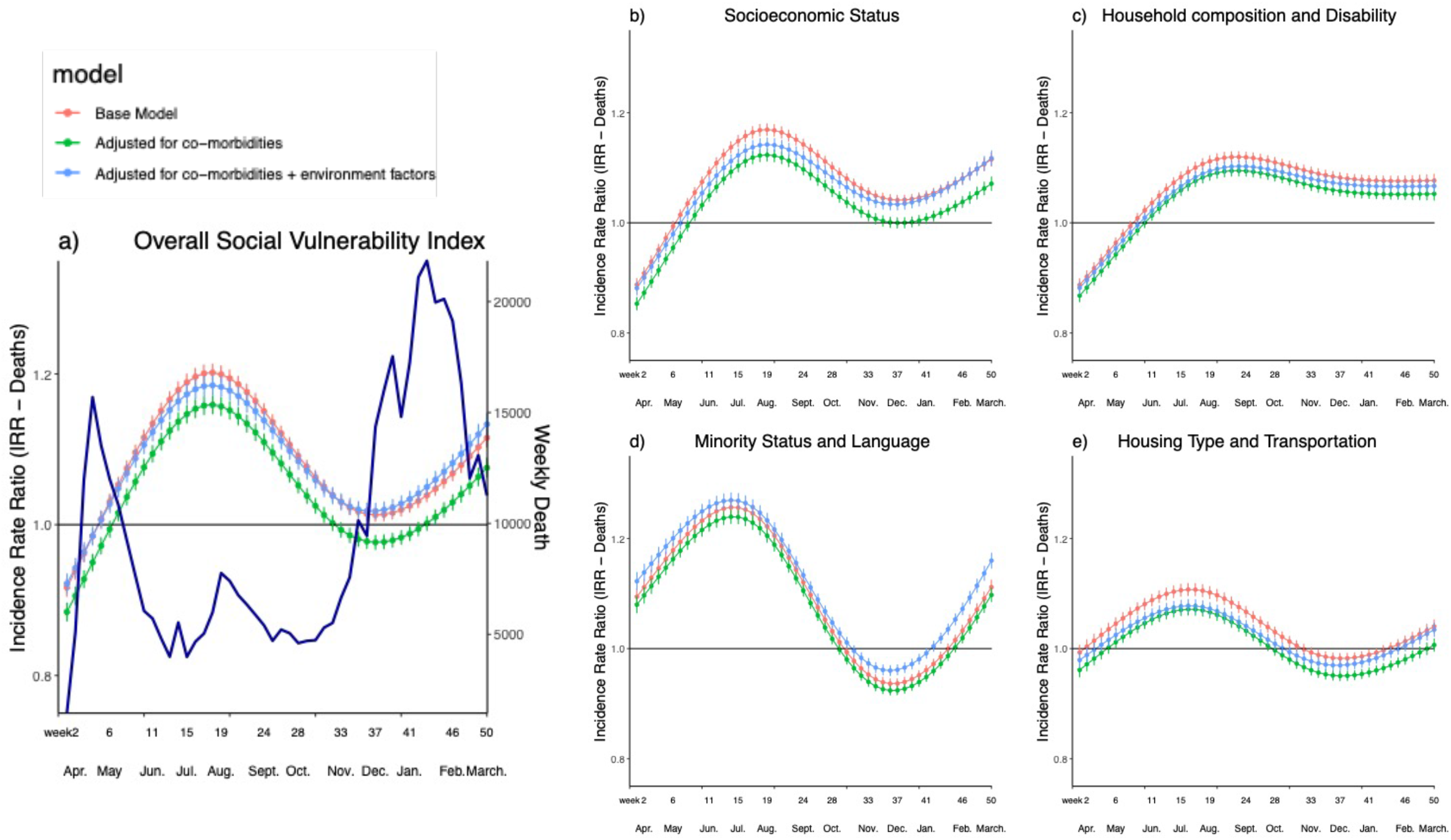
Temporal association between COVID-19 death per capita and the a) county-level Social Vulnerability Index (SVI) and its subcomponents: b) Socioeconomic Status, c) Household Composition and Disability, d) Minority Status and Language, and e) Housing Type and Transportation between March 22^nd^, 2020 and March 6^th^, 2021. The base model (red lines) is adjusted for proportion of population age ≥65 years and state-level COVID-19 testing. The green lines are additionally adjusted for CMS average Hierarchical Condition Category (HCC) score (proxy for comorbidities). The blue lines are additionally adjusted for environmental factors including average daily temperature (degrees Fahrenheit), average daily precipitation, and average particulate matter of diameter ≥ 2.5 micrometers (PM_2.5_). Of note, proportion age ≥65 years not included as a covariate for models for overall social vulnerability index and household composition/disability because these indices contain the age variable.

#### SVI Subcomponents (Figure 2)

The SVI subcomponent of minority status and language was significantly and positively associated with COVID-19 death per capita from the beginning of the analysis. The strength of the association decreased starting in late-July, 2020 (Week 17), and became negatively-associated starting in mid October, 2020 (Week 30) when the third peak in deaths were observed. It became positively associated again starting in early January 2021 (Week 41), with a decrease in deaths.

The SVI subcomponent of socioeconomic status was independently and positively associated with death per capita after accounting for co-morbidities and environmental factors starting in late May, 2020 (Week 9). While the strength of the association fluctuated throughout the duration of the pandemic, it remained a positive predictor and became more strongly associated with the death per capita starting early January 2021 (Week 41).

The index of county-level household composition and disability became positively associated with death per capita in early June 2020 (Week 11) and remained associated throughout the duration of the pandemic. Housing type and transportation became positively associated starting mid-May 2020 (Week 7), with the association weakening around early August (Week 19) and diminishing by early October 2020 (Week 28), but it became positively associated with death rate around February 2021 (Week 46). Of note, in analysis additionally adjusting for percentage of residents under the federal poverty limit, for SVI subthemes of minority status and language, household composition and disability, and housing type and transport, similar trends are noted (Supplemental Figure 2).

### Overall and Temporal Associations between race/ethnicity and COVID-19 Incidence & Death per Capita

In order to further investigate the association of minority status with worse COVID-19 outcomes, we compared the rate of infections and death per capita according to the proportion of whites, Hispanics and blacks within each county, based on county level data from 2015-2019 from the U.S. Census Bureau ACS database.^4^ Proportionately more blacks reside in the southern U.S. and Hispanics in southwestern states. Cumulatively for the full year analysis, county-level increase in proportion of Black residents (adjusted IRR per 10% increase: 0.99, 95% CI [0.98, 1.00], p=0.01) was associated with lower, while increase in proportion of Hispanic residents (adjusted IRR per 10% increase 1.07, 95% CI [1.05, 1.08], p<0.001) was associated with higher COVID-19 incidence after adjusting for comorbidities using the HCC score and environmental factors (**Table 3**). While county-level increase in proportion of Black residents (adjusted IRR per 10% increase: 1.00, 95% CI [0.99,1.02], p=0.85) was not associated with COVID-19 death per capita, county-level increase in Hispanic residents (adjusted RR per 10% increase: 1.07, 95% CI [1.05, 1.08], p<0.001) was independently associated with higher COVID-19 death per capita (**Table 3**).

**Table 3.**
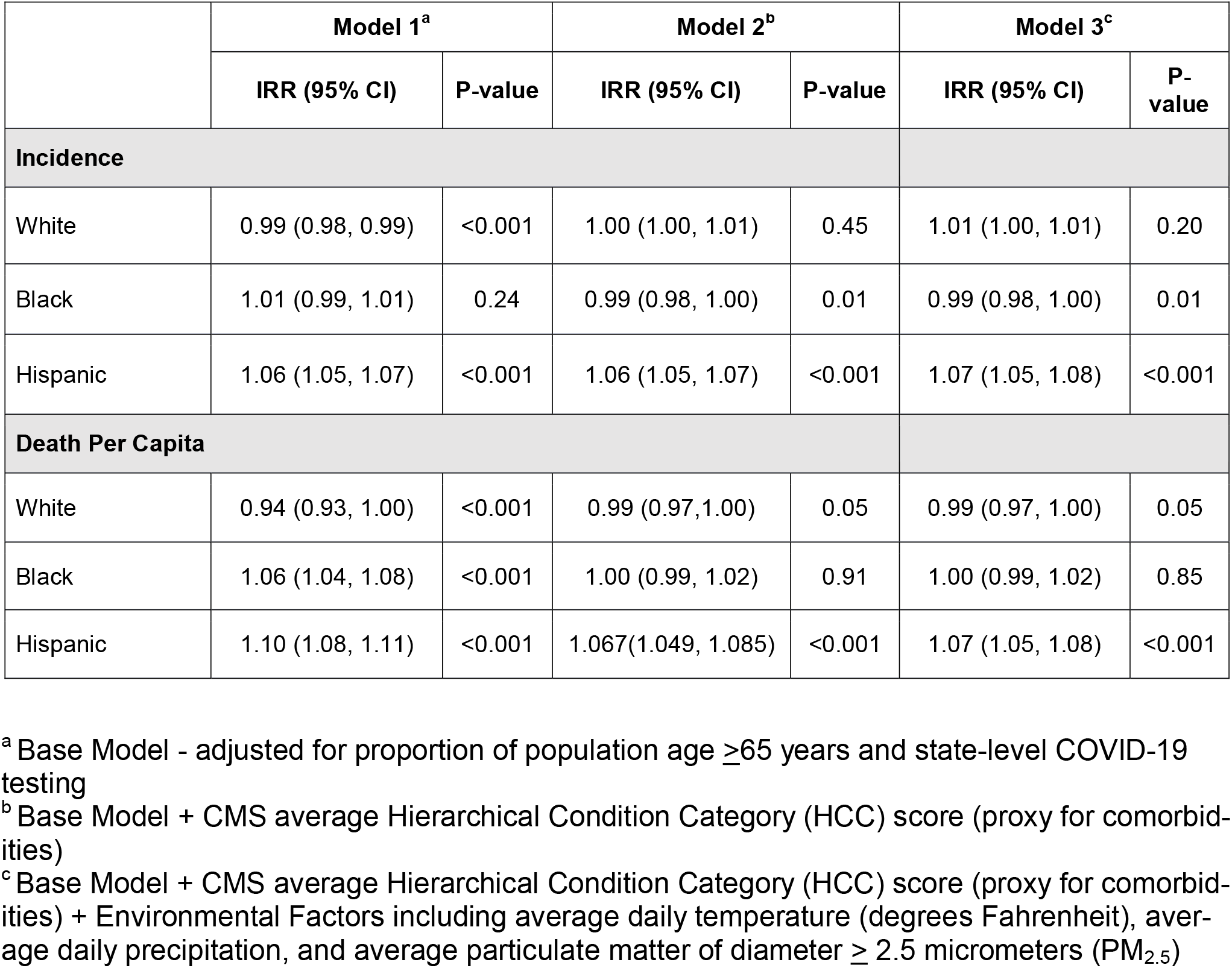
Overall association of county-level race/ethnic composition (Incidence Risk Ratio [IRR] per 10% increase) with incidence and case fatality rate of COVID-19 as of March 06, 2021

#### Temporal Trends

Counties with a greater proportion of Black residents had the highest incidence of infection and death per capita from the start of the study period till about mid to late August 2020 (Week 22) (**Figure 3**). For a period of 10 weeks between early November 2020 and early January (for incidence) and 14 weeks between early November 2020 and mid-January 2021 (for death per capita), counties with greater proportion of Black residents had lower incidence and death per capita. By January 2021 (Week 42), this trend reversed such that counties with higher proportion of Black residents again had worse outcomes. Out of the 50 weeks included in our analysis, counties with higher proportion of Black residents had higher incidence during 40 weeks (80% of the analysis time frame) and higher death per capita during 36 weeks (72% of the analysis time frame). Counties with higher proportion of White residents showed opposite trends. During a period of 8 weeks between November 2020 and early January 2020, counties with higher proportion of White residents had higher incidence, which coincided with higher overall cases in the U.S. (“third wave”). Similarly, for a period of 11 weeks between November 2020 and January 2020, these communities had higher death per capita.

**Figure 3:**
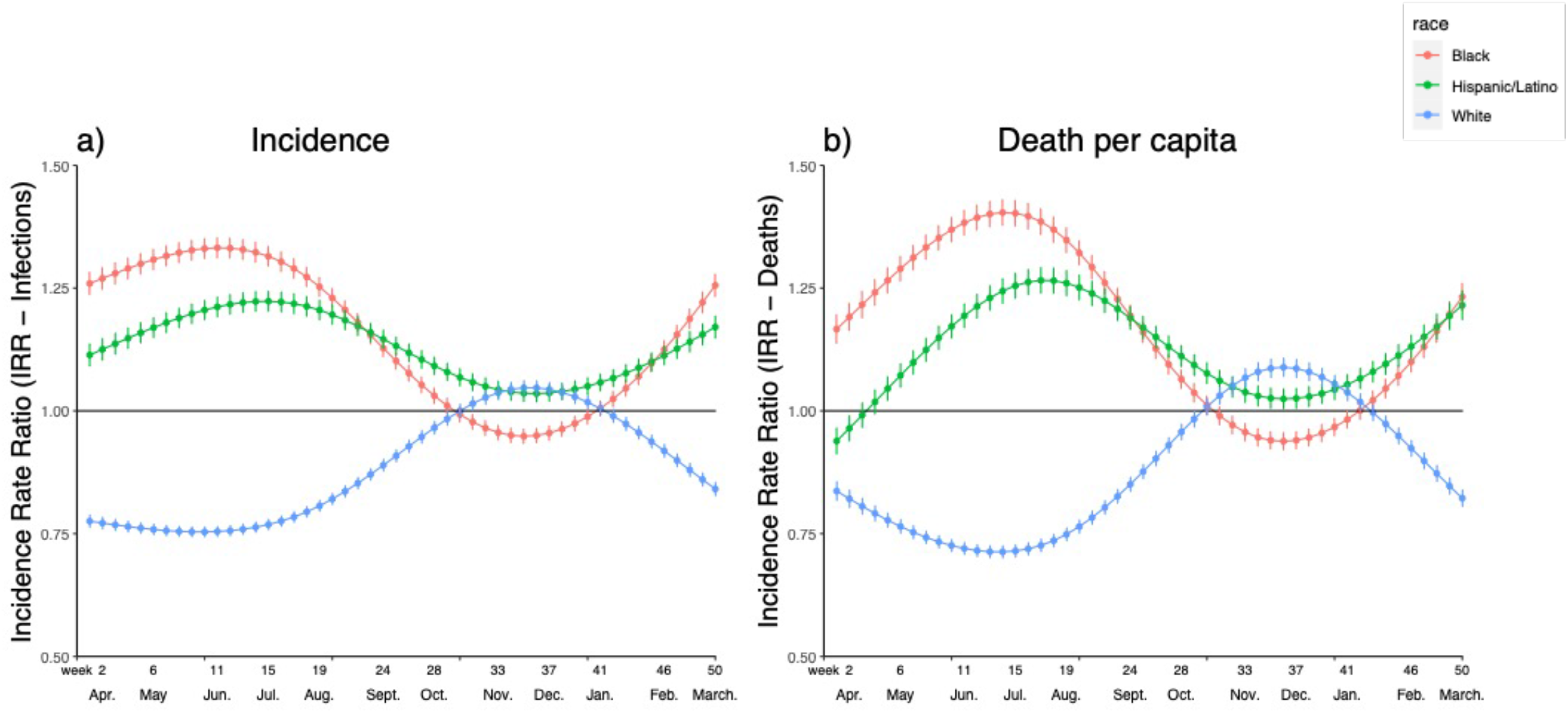
Temporal association of county-level racial composition (Black, Hispanic/Latino, White) and COVID-19 a) incidence and b) death per capita between March 22^nd^, 2020 and September 26^th^, 2020 after adjusting for proportion of population age ≥65 years, state-level COVID-19 testing, CMS average Hierarchical Condition Category (HCC) score (proxy for comorbidities) and environmental factors including average daily temperature (degrees Fahrenheit), average daily precipitation, and average particulate matter of diameter ≥ 2.5 micrometers (PM_2.5_).

Counties with higher proportion of Hispanic residents had both higher incidence and death per capita throughout the entire duration of the analysis.

When the geographical changes in the incidence and mortality rates from COVID-19 are examined over the year, it is apparent that whereas the early part of the pandemic affected the northeastern U.S. and areas in the south-east and southwestern U.S., areas that are enriched for minority populations, by the end of 2020 when the pandemic was at its peak (“third wave’), the mid-western states, with predominantly White populations, had the highest prevalence and mortality rates. By the Spring of 2021, the geographic distribution of cases again changed back to the areas affected initially during the pandemic **(Figure 4)**.

**Figure 4:**
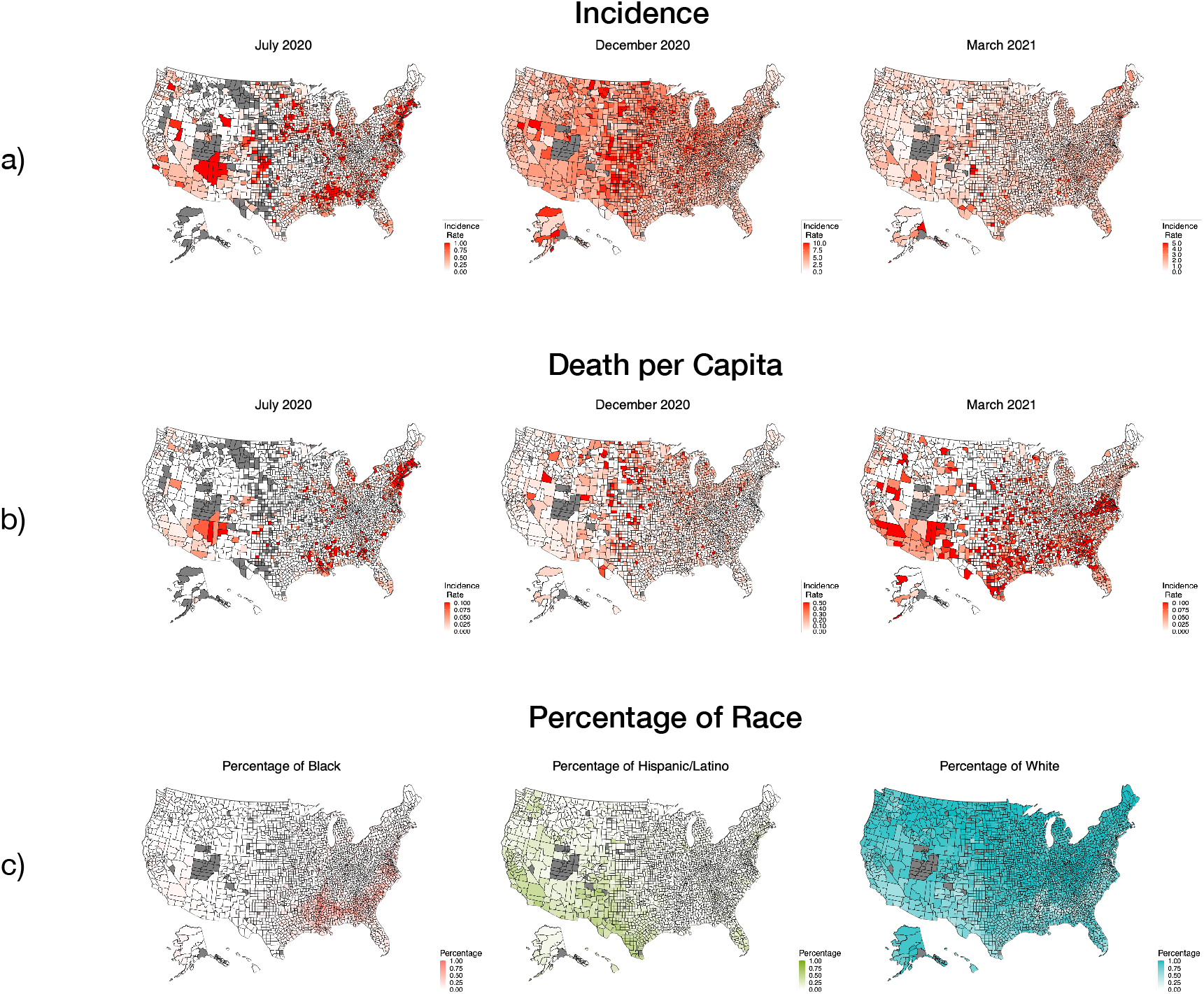
County-level map of the U.S. showing a) incidence and b) death per capita for COVID-19 across three timepoints – July 2020, December 2020, and March 2021. County-level proportion of Black, Hispanic/Latino, and White residents is shown in (c). As shown, Black and Hispanic residents are disproportionately represented in the southeast and southwestern US, where outcomes were worst in July 2020 and again in March 2021. Midwestern states where there are less diverse communities (higher proportion of White residents) showed worst outcomes in December 2020.

## Discussion

Herein, we demonstrate that U.S. counties with higher social vulnerability had an overall higher incidence of infection and death per capita from COVID-19. For every 10-percentile increase in SVI, indicative of increased social vulnerability, the incidence and death per capita of COVID-19 is 2% and 5% higher, respectively. However, there is a great deal of temporal variation in the association between SVI and COVID-19 outcomes, throughout the duration of the pandemic. SVI became an independent predictor of incidence of infections starting in April 2020, becoming an increasingly important predictor until August 2020. By late October, when the pandemic was at its third peak, SVI was no longer a predictor of the incidence of infections.

The absence of SVI being a predictor of COVID-19 outcomes coincided with a sharp increase in cases and deaths within the U.S. between early November 2020 and early January 2021, when counties with higher proportion of White residents were disproportionately represented in COVID-19 cases and deaths. Once the winter peak in cases reversed, higher SVI communities again began to experience worse COVID-19 outcomes.

While overall we demonstrate that socially vulnerable communities bear a disproportionate share of the burden of worse outcomes, during the time with the highest COVID-19 incidence and deaths (i.e the “third wave”) in the U.S., it is of great interest that SVI became a non-significant predictor of incidence and death. There are potentially several explanations for these trends and our analyses with the temporal associations between the subcomponents of the SVI are of special interest. Our analysis demonstrates that especially during the early phases of the pandemic, communities with a greater share of minority populations rather than socioeconomic disadvantage or crowding, were disproportionately bearing the disastrous effects of the pandemic. Our more in-depth analysis of racial composition data, beyond the SVI subcomponent of minority status and language, demonstrated nationwide trends that counties with a large share of Black and Hispanic residents had especially worse outcomes during the pandemic prior to the third wave between November 2020 to January 2021. During this period, majority white communities demonstrated the highest incidence and death rates. Our nationwide results are congruent with a more in-depth analysis completed in Cuyahoga County, OH and Wayne County, MI – both counties that are socially vulnerable, but Wayne County has a higher proportion of Black residents and thus, had worse COVID-19 outcomes including death and hospital utilization.^10^ In addition, these temporal changes may at least partly also be due to the geographical spread of COVID-19 infections in the U.S.. Whereas the pandemic affected the northeastern, southeastern and southwestern states during the early and late phases of the year of study, it was predominantly affecting the mid-western and central states, that are proportionately less diverse, during the third peak observed in the Winter months of 2020. While we controlled for community-level comorbidity burden, communities with a higher proportion of minority populations are vulnerable to worse health outcomes due to other factors above and beyond what is measured in the SVI, including structural racism, marginalization, and poor healthcare access.^23^ These factors need to be further studied.

Shortly following re-openings across the country in late May 2020, the socioeconomic component of the SVI became an independent predictor of worse COVID-19 outcomes and follows a similar trend to the overall SVI throughout the duration of our analysis. There is emerging evidence using cell phone data demonstrating that low income communities have been less able to socially distance during the COVID-19 pandemic, likely due to a multitude of factors including less capacity to work from home, or to take paid or unpaid time off from work, and limited savings.^24,25^ During the COVID-19 pandemic, data has suggested that Hispanic communities in the U.S. are particularly vulnerable to financial insecurities compared to other racial/ethnic groups due to their disproportionate representation in industries that have been most affected by the pandemic and having jobs that cannot be performed from home.^26^ We notice that temporal trends in incidence and death per capita for communities with greater proportion of Hispanic residents closely mirror that of the socioeconomic component of the SVI and thus, low SES may partially explain why Hispanic communities have had the worst overall COVID-19 outcomes for the duration of our analysis.

Another potential mechanism may be that low socioeconomic status communities have a higher burden of preexisting health conditions^24^, but our findings are independent of community-level comorbidities. Finally, lower income communities are also more likely to live in multi-family crowded environments^27^ and studies completed in restricted geographic locales such as New York City show the importance of these vulnerability markers^28^ in COVID-19 outcomes. In accordance with these studies, we are seeing similar associations with both household composition and disability and housing type and transportation with COVID-19 outcomes throughout the duration of the analysis time period. ^24,25^ However, future studies using individual patient level information across the U.S. need to be conducted to further clarify these associations.

## Limitations

One of the major limitations of our study is that is mainly descriptive, ecological, and uses only county-level data, which does not allow us to account for individual characteristics that may drive COVID-19 outcomes in socially vulnerable communities. In addition, we used data collected from different data sources, each of which was gathered at slightly varying time points and as such, may not completely represent the features of the community at the time of our analysis. We attempted to account for as many confounders as possible but recognize that we may not have been able to adjust for all confounders (including vaccinations) driving the associations seen in this analysis. In terms controlling for county-level comorbidity, we use the HCC risk score, which was designed to reflect healthcare access and hospital admissions in a geographic area, as a proxy which may impact the associations seen. Therefore, studies incorporating individual patient level data which includes more confounders are needed to further delineate associations seen in this ecological study. Finally, we focused our analysis on county-level proportion of Black and Hispanic residents within the U.S. and do not extend it to include Asian or Native American residents. Future studies that encompass other minority groups and examine trends presented in our study on a worldwide basis are needed.

## Data Availability

All data used is publicly available. List of data sources is listed in the supplementary materials

## Author contributions

Shabatun J. Islam, MD - Concept, drafting and editing of manuscript, statistical analysis

Aditi Nayak, MD - Concept, drafting and editing of manuscript

Yingtian Hu, BS – Statistical analysis, review of manuscript

Anurag Mehta, MD- Concept, drafting and editing of manuscript

Katherine Dieppa, M.Eng. – Statistical analysis, review of manuscript

Zakaria Almuwaqqat, MD, MPH - Statistical analysis, review of manuscript

Yi-An Ko, PhD - Statistical analysis, review of manuscript

Shivani A. Patel, MPH, PhD - Concept, review of manuscript

Abhinav Goyal, MD, MHS - Concept, review of manuscript

Samaah Sullivan, PhD - Concept, review of manuscript

Tené T. Lewis, PhD - Concept, drafting and editing of manuscript

Viola Vaccarino, MD, PhD - Concept, drafting and editing of manuscript

Alanna A. Morris, MD, MSc - Concept, drafting and editing of manuscript

Arshed A. Quyyumi, MD - Concept, drafting and editing of manuscript, review of manuscript

