## Supplementary Material for "Ecological Analysis of the Temporal Trends in the Association of Social Vulnerability and Race/Ethnicity with County-Level COVID-19 Incidence and Outcomes in the United States"

**Table S1.** Data sources used in the analysis (publicly available)

| **Data** | **Source** | |
| --- | --- | --- |
| **Outcomes** | | |
| **Case Fatality Rate** | Johns Hopkins Center for Systems Science and Engineering database, accessed May 28, 2021: https://coronavirus.jhu.edu/us-map | |
| **Incidence** | Johns Hopkins Center for Systems Science and Engineering database, accessed May 28, 2021: https://coronavirus.jhu.edu/us-map | |
| **Exposures** | | |
| **Social Vulnerability Index** | Centers for Disease Control and Prevention (CDC) Geospatial Research, Analysis, and Services Program (GRASP) database, accessed May 28, 2021: https://svi.cdc.gov/ | |
| **Racial Composition** | US Census Bureau, accessed May 28, 2021: https://www.census.gov/data.html | |
| **Confounders** | | |
| **Proportion age ≥65 years** | Centers for Disease Control and Prevention (CDC) Geospatial Research, Analysis, and Services Program (GRASP) database, accessed May 28, 2021: https://svi.cdc.gov/ | |
| **Hierarchical Condition Category score** | Centers for Medicare and Medicaid Services (CMS), accessed May 28, 2021: https://www.cms.gov/Research-Statistics-Data-and-Systems/Statistics-Trends-and-Reports/Medicare-Geographic-Variation | |
| **Tests administered per state** | The COVID Tracking Project, accessed May 28, 2021: https://covidtracking.com/ | |
| **Environmental Factors** | Temperature | National Centers for Environmental Information, accessed May 28, 2021: https://www.ncei.noaa.gov/ |
|  | Precipitation | National Centers for Environmental Information, accessed May 28, 2021: https://www.ncei.noaa.gov/ |
|  | PM_2.5_ | Robert Wood Johnson Foundation, accessed May 28, 2021: https://www.countyhealthrankings.org/explore-health-rankings/rankings-data-documentation |

**Supplemental Figure 1:** Correlation plot of county-level overall SVI, its subcomponents, and county-level proportion of White, Black and Hispanic/Latino residents

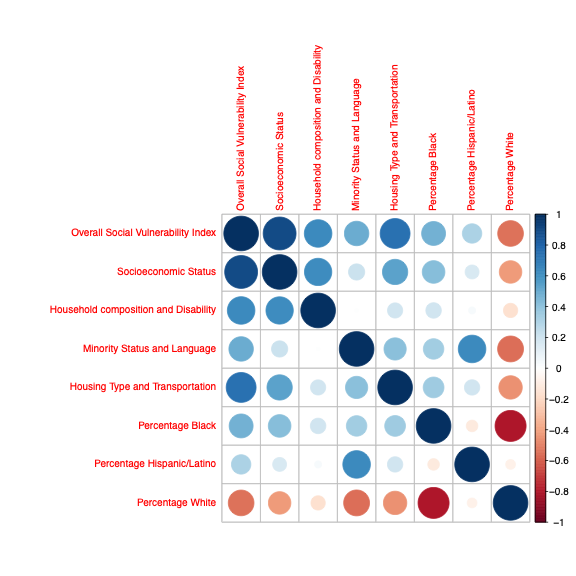

**Supplemental Figure 2:** Temporal association between COVID-19 incidence (top) and death (bottom) and SVI subcomponents of a) Household Composition and Disability, b) Minority Status and Language, and c) Housing Type and Transportation between March 22^nd^, 2020 and March 66^th^, 2020. The models labelled *adjusted for environmental factor* were adjusted for proportion of population age ≥65 years, state-level COVID-19 testing, CMS average Hierarchical Condition Category (HCC) score (proxy for comorbidities), and environmental factors including average daily temperature (degrees Fahrenheit), average daily precipitation, and average particulate matter of diameter ≥ 2.5 micrometers (PM_2.5_). Of note, proportion age ≥65 years not included as
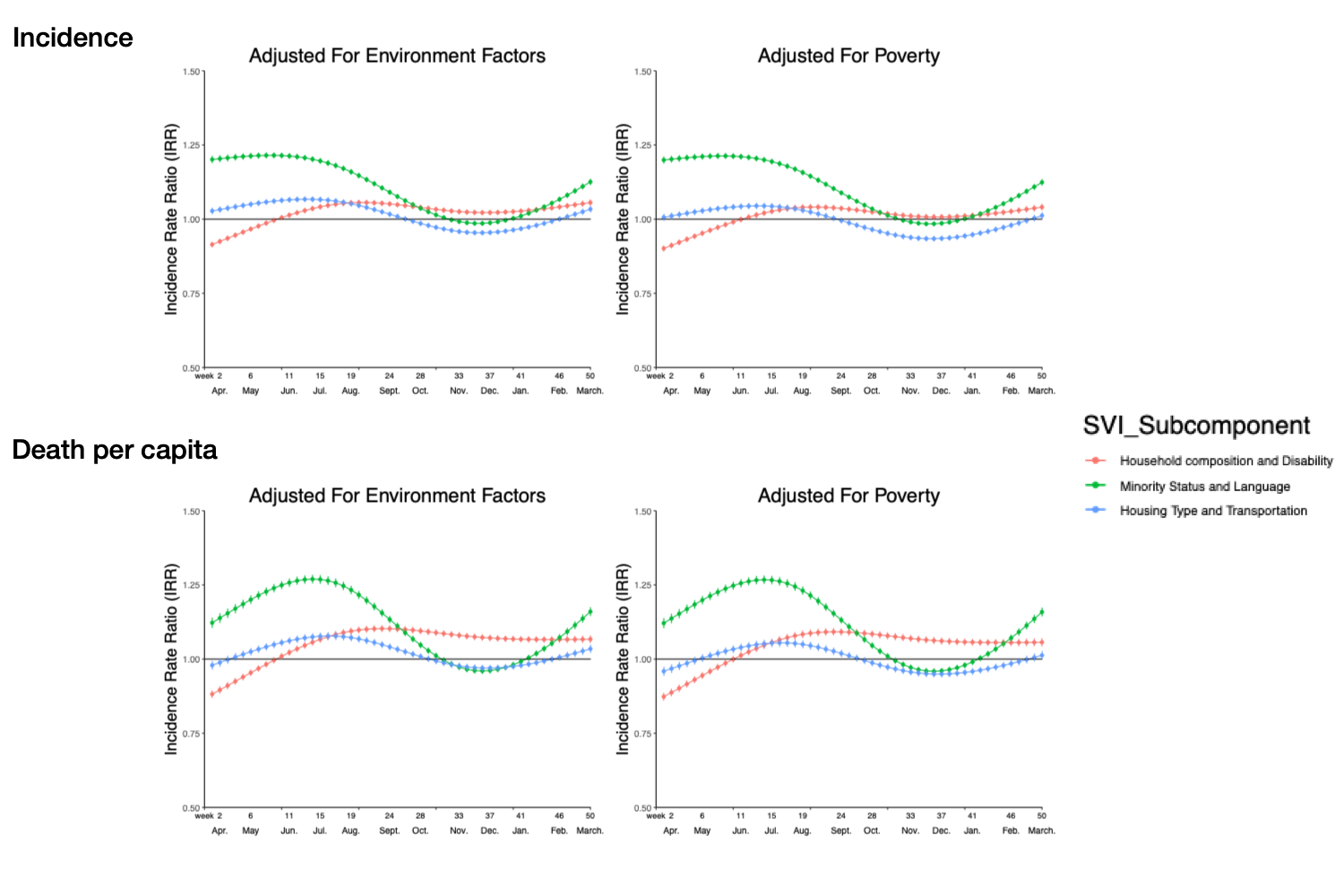
a covariate for household composition/disability because this index contains the age variable.
